# Association between Triglyceride-Glucose Index and Clinical Outcome in Patients with Acute Decompensated Heart Failure with Preserved Ejection Fraction

**DOI:** 10.1101/2025.05.13.25327566

**Authors:** Katsuomi Iwakura, Atsunori Okamura, Yoshitaka Iwanaga, Yasushi Koyama, Nobuaki Tanaka, Masato Okada, Masahiro Seo, Takahisa Yamada, Masamichi Yano, Takaharu Hayashi, Yoshio Yasumura, Yusuke Nakagawa, Shunsuke Tamaki, Akito Nakagawa, Katsuki Okada, Yohei Sotomi, Shungo Hikoso, Daisaku Nakatani, Yasushi Sakata, PURUSUIT-HFpEF investigators

## Abstract

**Background:** Although insulin resistance (IR) is associated with a higher risk of incident heart failure (HF), it is not fully understood whether IR could affect clinical outcomes in patients with established HF. We investigated the relationship between the triglyceride-glucose (TyG) index, a simple surrogate marker for IR, and clinical outcomes in patients with HF with preserved ejection fraction (HFpEF).

**Methods:** This retrospective analysis from the PURSUIT-HFpEF registry included 917 patients hospitalized for decompensated HFpEF. The TyG index was calculated at discharge as ln(triglyceride [mg/dL] × fasting blood glucose [mg/dL]/2). The primary outcomes were all-cause death and major adverse cardiovascular events (MACEs; a composite of all-cause death, heart failure hospitalization, and stroke).

**Results:** The median age of patients was 83 years, 44.7% was male, and 39.2% had diabetes. The median BMI was 21.5 kg/m², with 20.9% having BMI <18.5 kg/m ². During a median follow-up of 387 days, 168 deaths and 343 MACEs occurred. A stepwise Cox hazard model revealed that higher TyG index was independently associated with lower risk of all-cause death (HR 0.53, 95% CI 0.38-0.75) and MACEs (HR 0.77, 95% CI 0.61-0.97). When patients were divided into quartiles based on TyG index, the incidences of both outcomes were significantly lower in higher TyG quartiles (all-cause death; p=0.0003, MACEs; p=0.007 by log-rank).

**Conclusions:** In this predominantly elderly, low BMI cohort with established HFpEF, higher TyG index was paradoxically associated with better clinical outcomes. These findings imply a complex relationship between IR and HFpEF outcomes.

**Clinical Perspective:** *What is new?:*

- The triglyceride-glucose (TyG) index is a novel, easily calculated surrogate marker for insulin resistance (IR) using only serum triglyceride and fasting blood glucose levels.
- A higher TyG index, reflecting greater IR, was paradoxically associated with a lower incidence of all-cause mortality and major adverse cardiovascular events in patients hospitalized with heart failure with preserved ejection fraction (HFpEF) over approximately one year of follow-up.
- This association remained consistent across various subgroups stratified by age, sex, BMI, diabetes status, and other risk factors.

*What are the clinical implications?:*

- A paradoxical relationship between IR and clinical outcomes may exist in predominantly older, low-BMI Japanese patients with established HFpEF.
- The prognostic impact of IR in established heart failure appears complex and may vary depending on patient characteristics.

---

Insulin resistance (IR) plays a major role in the development of cardio-kidney-metabolic syndrome^1^. It is not only related with development of atherosclerotic cardiovascular disease (ASCVD) but also with incident heart failure (HF) in both in diabetic- and non-diabetic patients ^2–4^. A meta-analysis of prospective studies using the homeostasis model assessment for insulin resistance (HOMA-IR) confirmed that higher level of IR is associated with a higher risk of incident HF after adjustment of traditional risk factors^5^. IR is frequently observed in non-diabetic patients both with HF with reduced ejection fraction (HFrEF) and HF with preserved ejection fraction (HFpEF)^6,7^, and those with HFrEF show more severe IR than those with HFpEF^7,8^. However, it is not fully elucidated whether IR could affect clinical outcomes in patients with established HF.

Whereas the glucose clamp method is the gold standard to evaluate IR, it is expensive and time-consuming for daily clinical practice or for large scale clinical studies. Surrogate markers such as HOMA-IR are widely used for clinical assessment of IR. The triglyceride-glucose (TyG) index was a novel surrogate marker for IR which is easily calculated using only serum triglyceride and fasting blood glucose, and is well correlated with IR measured with glucose clamp method^9^. The TyG index demonstrated higher predictive power for both prevalent and incident metabolic syndrome compared to HOMA-IR^10,11^ It also showed stronger associations with arterial stiffness and with coronary artery calcification than HOMA-IR^12^. IR assessed by TyG index was well associated with the incidence of HF^13,14^. In the present study, we investigated the relationship between the TyG index and clinical outcomes in patients hospitalized for acute decompensation HFpEF.

## Methods

### Study Population

We retrospectively analyzed the data from the Prospective Multicenter Observational Study of Patients with Heart Failure with Preserved Ejection Fraction (PURSUIT-HFpEF), a multicenter, observational registration study enrolling consecutive patients hospitalized for acute decompensated HFpEF (left ventricular ejection fraction (LVEF) ≥ 50%) in 31 collaborating hospitals [UMIN-CTR ID: UMIN000021831]. Details of the entry criteria and data collection have been described elsewhere^15^. Briefly, patients admitted with acutely decompensated HFpEF were registered, and their clinical data including medications, laboratory tests, and ECG were collected on admission, at discharge, and 1 year after discharge. Acutely decompensated HFpEF was diagnosed based on the following criteria: (1) clinical symptoms and signs according to the Framingham Heart Study criteria, (2) LVEF on admission ≥50%, and (3) serum N-terminal pro-B type natriuretic peptide (NT-proBNP) ≥400 pg/mL or brain natriuretic peptide ≥100 pg/mL. T2DM was diagnosed based on clinical history, or on fasting plasma glucose and/or hemoglobin A1c (HbA1c) during hospitalization based on Japanese Clinical Practice Guideline for Diabetes 2019. Oral glucose tolerance test was not mandatory for the present study.

This study was conducted in accordance with the Declaration of Helsinki. The study protocol was approved by each corresponding hospital’s Ethics Committee. Informed consent was obtained from each patient by one of the investigators before the study.

### Data Collection

We collected patient data including risk factors and history of major comorbidities such as DM, hypertension, dyslipidemia, smoking, chronic kidney disease, history of HF hospitalization, prior myocardial infarction, prior stroke and malignancy, and history of percutaneous coronary intervention or coronary artery bypass graft. Blood tests, standard 12-lead ECG recording, chest radiography, and echocardiography were performed immediately after admission, before discharge, and 1year after discharge. TyG index was calculated as ln (serum triglyceride [mg/dL] x fasting blood glucose [mg/dL]/2), using blood test data at the time of discharge.

Echocardiography was performed before discharge in each hospital. LVEF was determined by the modified Simpson’s technique using images from the apical 2- and 4-chamber view. The early diastolic mitral annular velocity (e’) was measured using tissue Doppler imaging from the septal and lateral portion of the mitral annulus in the apical four-chamber view, and the average of the septal and lateral e’ velocities were calculated. E/e’ ratio was determined as the ratio of mitral E velocity to average e’ velocity. Left atrial volume was determined by the modified Simpson’s technique using images from the apical 2 view, and it was indexed by body surface area as left atrial volume index (LAVI). Left ventricular diastolic dysfunction was determined according to the 2016 American Society of Echocardiography guidelines, using LAVI, e’ velocity E/e’ ratio and peak tricuspid regurgitation velocity.

Study patients were followed up by direct contact or telephone interview to observe all-cause death or major adverse cardiovascular events (MACE), which was defined as a composite of all-cause death, HF hospitalization, and stroke. For patients whose survival information could not be determined by these means, the data from National Vital Statistics of Japan, which includes all death records in Japan reported by prefectural public health centers, were used with the permission of the Ministry of Health, Labor and Welfare in accordance with the Statistics Act in Japan.

### Statistical Analysis

The normality of the continuous variables was assessed using the Kolmogorov-Smirnov test. Normally distributed variables are presented as the mean ± standard deviation and non-normally distributed variables as the median (interquartile range). We made comparisons by the Student’s T test or one-way ANOVA for normally distributed variables, and by the Mann-Whitney U test or the Kruskal-Wallis test for non-normally distributed variables. Statistical significance for comparisons among more than three groups was assessed using the Bonferroni correction. Categorical variables were compared with Fisher’s exact test, and the p-value was determined using a simulation approach because of the substantial amount of data. Spearman’s rank correlation was performed to analyze the correlation between TyG index and continuous values. A stepwise Cox proportional hazard model for all-cause death or for MACEs was constructed including TyG index and the relevant parameters such as age, sex, body mass index (BMI), history of diabetes, hypertension, dyslipidemia and atrial fibrillation, systolic and diastolic blood pressure at discharge, and blood test results (c-reactive protein (CRP), estimated glomerular filtration ratio (eGFR), fasting blood glucose hemoglobin A1c, triglyceride, hemoglobin concentration, high density lipoprotein cholesterol (HDL-c), low-density lipoprotein cholesterol (LDL-c), uric acid, and NT-proBNP) at discharge. Event-free survival analysis was performed using the Kaplan-Meier method with the log-rank test for group comparisons. All statistical analyses were performed using R (R Foundation for Statistical Computing, Vienna, Austria) and R with a graphical user interface EZR (Saitama Medical Centre, Jichi Medical University, Japan).

## Results

### Patients Characteristics

Among 1095 patients hospitalized for decompensated HFpEF and registered in PURSUIT-HFpEF between June 2016 and December 2021, we calculated the TyG index at discharge in 917 patients (83.7%). The TyG index was not obtained in remaining 178 patients due to death during hospital stay (19 patients) or to missing data at discharge (159 patients). The median age was 83 (77 - 87) years and 729 patients (79.5%) were older than 75 years. Male patients consisted of 410 (44.7%) patients. Hypertension was observed in 774 patients (84.4%) and dyslipidemia in 521 patients (56.8%) (Table 1). Diabetes was observed in 360 patients (39.2%), and they were younger (82 (76 - 86) years vs. 83 (77 - 88) years, p<0.001), more male (48.9% vs 42.0%, p=0.04), and had higher incidence of hypertension (89.4% vs. 81.3%, p<0.001) and of dyslipidemia (56.6% vs.33.8%, p<0.001) than those without diabetes. Hemoglobin A1c in diabetic patients was 6.7 (6.3 – 7.2) % and that in non-diabetic patients was 5.7 (5.5 -6.0) % (p<0.0001). Median BMI was 21.5 (19.0 – 24.3) kg/m^2^, and 192 patients (20.9%) had BMI<18.5 kg/m^2^ and 185 patients (20.2) had BMI≥25.0 kg/m^2^. Diabetic patients had higher BMI (22.6 (19.9 – 25.7) kg/m^2^) than non-diabetic patients (20.8 (18.4 – 23.7) kg/m^2^, p<0.001). Figure 1 presented the distribution of Age, BMI and HbA1c in the study cohort.

**Figure 1:**
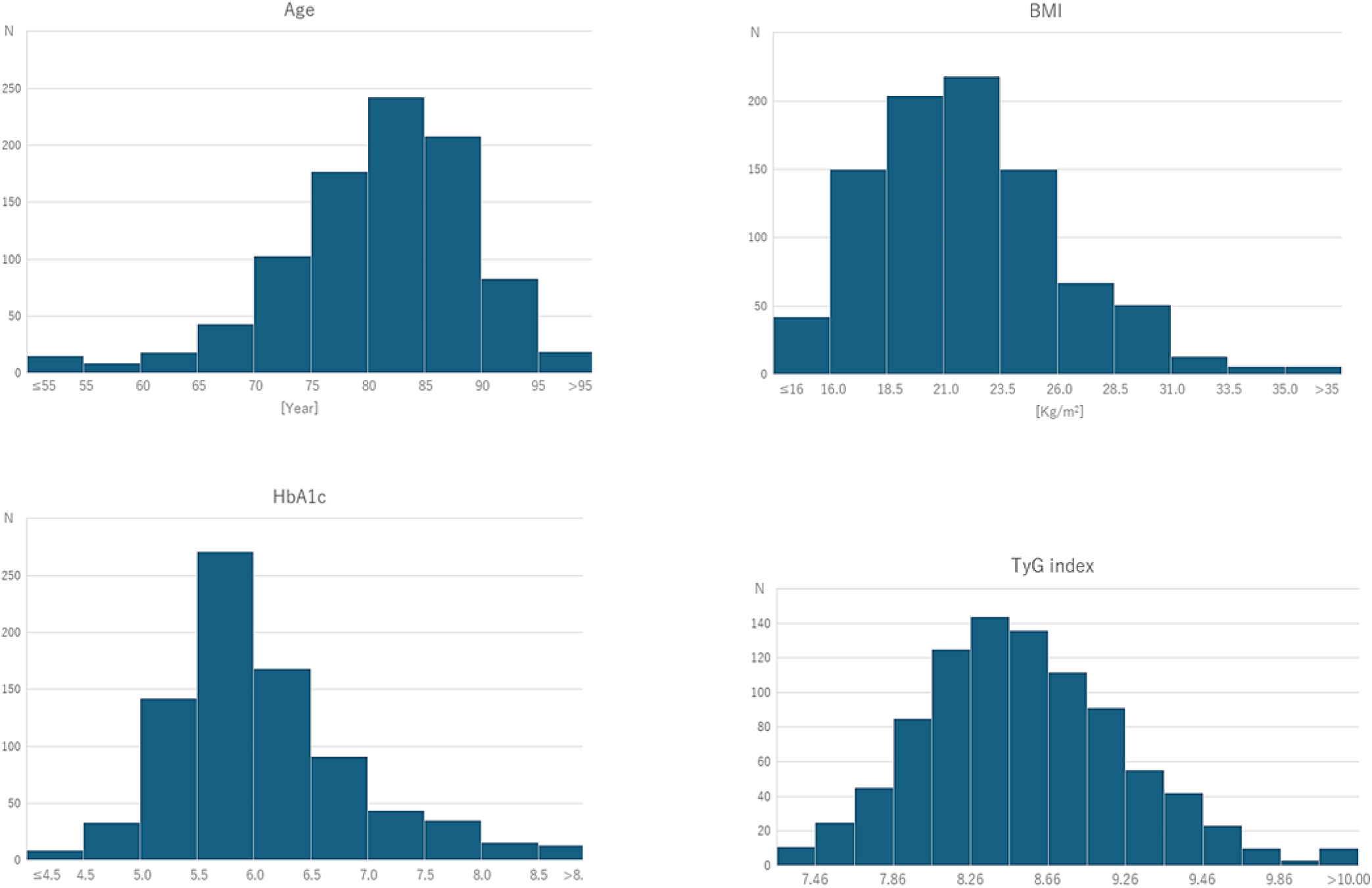
Distribution of age, BMI, HbA1c and TyG index in study patients. Distribution of age, BMI, HbA1c and TyG index in 917 patients with heart failure with HFpEF. The median age was 83 years and 729 patients (79.5%) was over 75 years (Upper left). The median value of BMI was 21.5 kg/m^2^ and 192 patients (20.9%) had BMI<18.5 kg/m^2^ while 185 patients (20.2) had BMI≥25.0 kg/m^2^ (Upper right). The TyG index followed normal distribution whereas age, BMI and HbA1c did not.

**Table 1.** Patients Characteristics.

|  | All patients | Q1<br>(7.26 to 8.15) | Q2<br>(8.16 to 8.50) | Q3<br>(8.51 to 8.87) | Q4<br>(8.88 to 10.60) | p |
| --- | --- | --- | --- | --- | --- | --- |
| **Number** | 917 | 229 | 229 | 229 | 230 |  |
| **Age, year** | 83 (77 - 87) | 83 (79 - 88) | 84 (78 - 88) | 82 (77 - 86) | 81.5 (74 - 86) <sup>¶,\|\|</sup> | <0.001 |
| **Male, n(%)** | 410 (44.7) | 109 (47.6) | 109 (47.6) | 89 (38.9) | 103 (44.8) | 0.19 |
| **Body mass index, kg/m<sup>2</sup>** | 21.5 (19.0 - 24.3) | 20.5 (17.9 - 22.6) | 21.2 (18.6 - 23.8) | 22.1 (19.5 - 24.7) <sup>¶,§</sup> | 23.2 (20.2 - 26.6) <sup>¶, **,††</sup> | <0.001 |
| **Hypertension, n(%)** | 774(84.4) | 183 (79.9) | 197 (86.0) | 189 (82.5) | 205 (89.1) <sup>*</sup> | 0.02 |
| **Dyslipidemia, n(%)** | 521 (56.8) | 59(25.7) | 97 (42.3) <sup>¶</sup> | 112 (48.9) <sup>¶</sup> | 124 (53.9) <sup>¶</sup> | <0.001 |
| **Diabetes, n(%)** | 360 (39.2) | 44 (19.2) | 73 (31.8) <sup>*</sup> | 96 (41.9) <sup>¶</sup> | 147 (63.9) <sup>¶, **,‡‡</sup> | <0.001 |
| **History of atrial fibrillation, n(%)** | 346 (37.7) | 102 (44.5) | 88 (38.4) | 86 (37.6) | 70 (30.4) <sup>*</sup> | 0.02 |
| **Systolic blood pressure, mmHg** | 119 (107 - 131) | 114 (104 - 127) | 118 (196 - 130) | 120 (107 - 131) <sup>*</sup> | 122 (108 - 133) <sup>¶</sup> | <0.001 |
| **Diastolic blood pressure, mmHg** | 65 (58 - 73) | 63 (57 - 71) | 65 (58 - 75) | 66 (59 - 74) | 65 (58 - 73) | 0.14 |
| **Blood glucose, mg/dL** | 98 (88 - 118) | 88 (81 - 95) | 94 (86 - 105) <sup>¶</sup> | 102 (93 - 119) <sup>¶, **</sup> | 126 (105 - 128) <sup>¶, **,‡‡</sup> | <0.001 |
| **Hemoglobin A1c, %** | 6.0 (5.6 - 6.5) | 5.7 (5.5 - 6.0) | 5.9 (5.5 - 6.3) <sup>*</sup> | 6.0 (5.7 - 6.5) <sup>¶</sup> | 6.4 (5.9 - 7.1) <sup>¶, **,‡‡</sup> | <0.001 |
| **Blood nitrogen, mg/dl** | 25.0 (18.6 - 34.0) | 23.0 (18.0 - 30.0) | 25.0 (19.4 - 34.8) | 23.0 (17.0 - 32.0) | 27.0 (19.0 - 38.8) <sup>‡, ††</sup> | 0.001 |
| **Creatinine, mg/dl** | 1.1 (0.9 - 1.5) | 1.0 (0.9 - 1.3) | 1.1 (0.9 - 1.5) | 1.1 (0.8 - 1.5) | 1.2 (0.9 - 1.9) <sup>¶, ††</sup> | <0.001 |
| **Estimated GFR, ml/min/1.73m<sup>2</sup>** | 42.9±19.3 | 46.9±18.0 | 42.7±19.5 | 43,5±20.3 | 38.3±18.7 <sup>¶, ††</sup> | <0.001 |
| **Uric acid, mg/dl** | 6.6 (5.4 - 8.0) | 6.0 (4.9 - 7.4) | 6.9 (5.6 - 8.1) <sup>¶</sup> | 6.6 (5.8 - 8.0) <sup>†</sup> | 7.1 (5.8 - 8.3) <sup>¶</sup> | <0.001 |
| **Total cholesterol, mg/dl** | 158 (136 - 181) | 148 (126 - 168) | 153 (134 - 174) | 164 (142 - 187)<sup>¶,§</sup> | 166 (147 - 194)<sup>¶, **</sup> | <0.001 |
| **HDL-c, mg/dl** | 43 (36 - 52) | 48 (41 - 57) | 44 (37 - 53)<sup>*</sup> | 43 (36 - 50)<sup>¶</sup> | 37 (32 - 46)<sup>¶, **, ‡‡</sup> | <0.001 |
| **LDL-c, mg/dl** | 91 (73 - 111) | 81.5 (66 - 100) | 90 (74 - 107)<sup>†</sup> | 95 (78 - 115)<sup>¶</sup> | 98 (77 - 122)<sup>¶,§</sup> | <0.001 |
| **Triglyceride, mg/dl** | 96 (73 - 126) | 63 (53 - 70) | 89 (78 - 98)<sup>¶</sup> | 112 (97 - 125)<sup>¶, **</sup> | 153 (127 - 189)<sup>¶, **, ‡‡</sup> | <0.001 |
| **Red blood cell count, x10<sup>4</sup>/μl** | 379±66 | 365±60 | 377±68 | 391±65<sup>¶</sup> | 382±69<sup>*</sup> | <0.001 |
| **Hemoglobin, g/l** | 11.3 (10.0 - 12.7) | 10.9 (9.7 - 12.2) | 11.2 (9.9 - 12.6) | 11.6 (10.3 - 13.0)<sup>‡</sup> | 11.4 (10.0 - 12.7) | 0.006 |
| **C-reactive protein, mg/l** | 0.27 (0.11 - 0.80) | 0.21 (0.09 - 0.82) | 0.3 (0.14 - 0.92) | 0.29 (0.12 - 0.80) | 0.33 (0.12 - 0.76) | 0.161 |
| **NT-proBNP, pg/ml** | 1110 (481 - 2380) | 1155 (575 - 2305) | 1150 (512 - 2430) | 1007 (500 - 2265) | 1101 (419 - 2618) | 0.748 |

\* p<0.05, † p<0.01, ‡ p<0.005, ¶ p<0.001 vs. Q1
§p<0.05, || p<0.01 \*\* p<0.001 vs. Q2
†† p<0.05, ‡‡ p<0.001 vs. Q3
Continuous variables are expressed as mean ± standard deviation or median (interquartile range). Categorical variables are expressed as number of patients (%). P value for the difference between subgroups.
GFR denotes glomerular filtration rate; HDL-c, high-density lipoprotein cholesterol; LDL-c, low-density lipoprotein cholesterol; NT-proBNP, N-terminal pro B-type natriuretic peptide.

### TyG Index

The mean value of TyG index in the 917 study patients was 8.54±0.53 (range 7.26 to 10.60). There was a significant correlation between TyG index and BMI (ρ= 0.26, p<0.001) or HbA1c (ρ= 0.34, p<0.001) (Figure 2). Patients with diabetes had higher TyG index than those without it (8.76±0.56 vs. 8.39±0.46, p<0.001).

**Figure 2:**
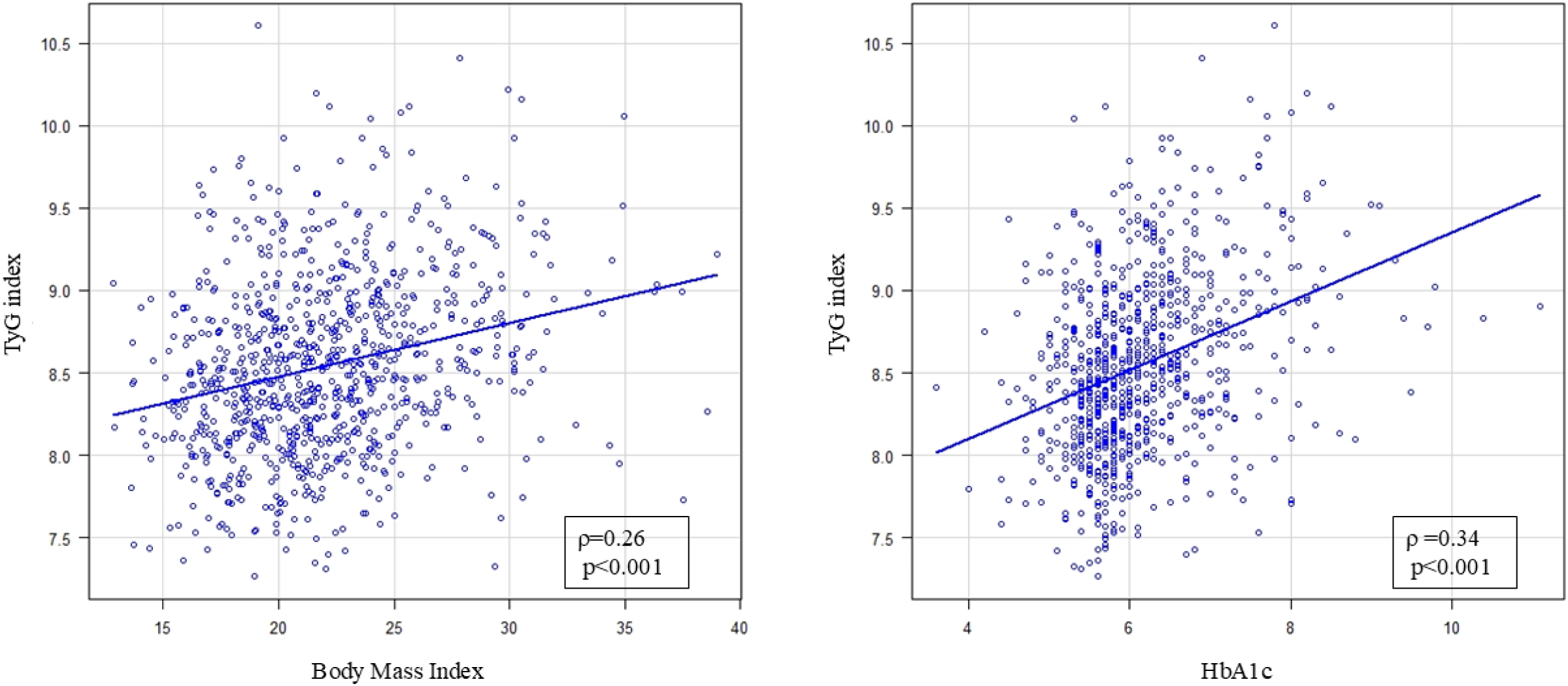
Correlation between TyG index and BMI or HbA1c. A significant correlation was observed between TyG index and body mass index (BMI) (ρ=0.26, p<0.001, Left) or HbA1c (ρ=0.34, p<0.001, Right).

We divided them into 4 groups based on the quartile of TyG index; Q1 (TyG index; 7.26 to 8.15), Q2 (8.16 to 8.50), Q3 (8.51 to 8.87), and Q4 (8.88 to 10.60). There were significant differences in age (p<0.001), BMI (p<0.001), incidences of hypertension (p=0.02), dyslipidemia (p<0.001), diabetes (<0.001), and history of atrial fibrillation (p=0.02) among 4 groups while no sex difference was observed(p=0.19) (Table 1). The Q4 group was significantly younger than the other 3 groups and no difference in age was observed among Q1, Q2 and Q3. The incidence of comorbidities was higher in the higher TyG groups (Table 1). Significant differences were observed between groups in blood test results at discharge (Table 1). The higher TyG groups had worse metabolic parameters and worse renal function, whereas they had higher blood cell count and hemoglobin concentration. Whereas no significant correlation was observed between TyG index and NT-proBNP (ρ=-0.0122, p=0.722), the higher TyG groups had higher NT-proBNP (p=0.046).

There was no significant difference in echo parameters before discharge except left ventricular end-diastolic diameter, LAVI and E/e’ ratio among 4 groups (Table 2). The Q1 group had a significantly lower E/e’ ratio than the Q3 (p=0.02) and Q4 group (p=0.006), whereas LAVI of the Q1 group was larger than that of the Q3 (p=0.01) and Q4 group (p=0.003). There was no difference in the prevalence of diastolic dysfunction among 4 groups (p=0.75, Table 2).

**Table 2.** Echocardiography parameters.

|  | All patients | Q1<br>(7.26 to 8.15) | Q2<br>(8.16 to 8.50) | Q3<br>(8.51 to 8.87) | Q4<br>(8.88 to 10.60) | p |
| --- | --- | --- | --- | --- | --- | --- |
| **Number** | 917 | 229 | 229 | 229 | 230 |  |
| **LV end-diastolic diameter, mm** | 46 (41 - 50) | 45 (40 - 50) | 45.0 (41.4 - 49.0) | 45.4 (41.0 - 50.0) | 46.6 (42.5 - 51.2) * | 0.02 |
| **LV end-systolic diameter, mm** | 29.2 (26.0 - 33.0) | 29.0 (25.7 - 32.0) | 29.0 (26.0 - 32.0) | 30.0 (26.0 - 33.0) | 30.0 (26.5 - 34.0) | 0.19 |
| **LV ejection fraction, %** | 61.0 (55.0 - 65.4) | 61.0 (55.0 - 65.7) | 61.0 (55.9 - 65.6) | 60.0 (55.0 - 64.6) | 61.4 (55.0 - 65.9) | 0.89 |
| **LV mass index, kg/m<sup>2</sup>** | 103.0 (85.4 - 124.5) | 101.8 (81.3 - 120.3) | 102.2 (85.7 - 123.4) | 102.5 (85.4 - 125.0) | 105.2 (86.6 - 128.6) | 0.26 |
| **Left atrial diameter, mm** | 44 (39 - 49) | 43 (39 - 50) | 44 (39 - 50) | 44 (39 - 49) | 45 (40 - 48) | 0.95 |
| **Left atrial volume index, ml/m<sup>2</sup>** | 50 (37 - 66) | 53 (39 - 72) | 51 (38 - 66) | 51 (36 - 66) * | 46 (35 - 60) ‡ | 0.005 |
| **E velocity, m/s** | 0.8 (0.6 - 1.0) | 0.8 (0.6 - 1.0) | 0.8 (0.6 - 1.0) | 0.8 (0.6 - 1.0) | 0.8 (0.6 - 1.0) | 0.94 |
| **E/A ratio** | 0.8 (0.6 - 1.2) | 0.8 (0.7 - 1.2) | 0.8 (0.6 - 1.3) | 0.8 (0.7 - 1.2) | 0.8 (0.6 - 1.1) | 0.97 |
| **E/e' ratio** | 12.5 (9.7 - 16.7) | 11.7 (8.9 - 15.2) | 12.3 (9.6 - 16.8) | 12.6 (9.8 - 18.2) * | 13.0 (10.4 - 17.5) † | 0.002 |
| **Diastolic wall strain,** | 0.28±0.12 | 0.27±0.13 | 0.29±0.11 | 0.27±0.13 | 0.27±0.11 | 0.51 |
| **Diastolic dysfunction** |  |  |  |  |  | 0.75 |
| **Normal diastolic function, n** |  | 64 (27.9) | 58 (25.3) | 57 (24.9) | 52 (22.6) |  |
| **Diastolic dysfunction, n** |  | 89 (38.9) | 101 (44.1) | 102 (44.5) | 97 (42.2) |  |
| **Indeterminate, n** |  | 76 (33.2) | 70 (30.6) | 70 (30.6) | 81 (35.2) |  |

\* p<0.05, † p<0.005, ‡ p<0.001 vs. Q1
Continuous variables are expressed as median (interquartile range). Categorical variables are expressed as number of patients (%). P value for the difference between subgroups. LV denotes left ventricle.

### TyG index and Clinical Outcomes

During follow up period 387 days (239 - 727 days), 168 death and 343 MACEs was observed. Stepwise Cox hazard model analysis selected TyG index as an independent predictor both for all-cause death and MACEs, as shown in Table 3. The hazard ratio (HR) of TyG index for all-cause death was 0.53 (95% confidence interval (CI); 0.38, 0.75, p<0.001) and that for MACEs was 0.77 (95% CI; 0.61, 0.97, p=0.02), indicating that TyG index was inversely correlated with all-cause death and MACEs. Kaplan-Meier survival curves indicated the difference in all-cause death (p=0.0003 by log-rank test) and MACEs (p=0.007 by log-rank test) among the 4 groups based on TyG index, and the highest TyG group (Q4) had lower incidence of all-cause death and MACEs (Figure 3). The Q3 and Q4 groups had significantly lower all-cause mortality than the Q1 group (p=0.004, Q1 vs Q3; p<0.001, Q1 vs. Q4) and lower rate of MACEs than the Q2 group (p=0.02, Q2 vs. Q3; p=0.04, Q2 vs. Q4).

**Figure 3.**
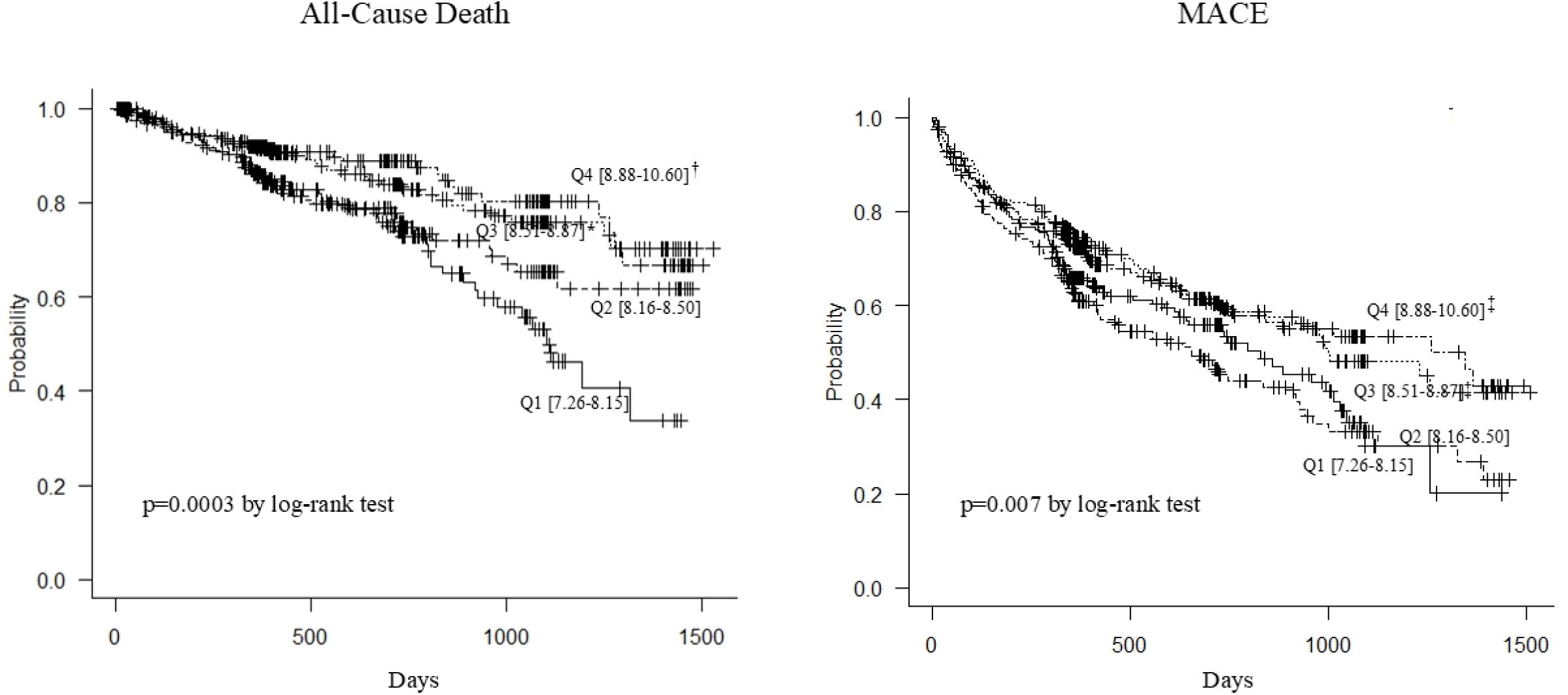
Kaplan‒Meier curves for MACEs and all-cause mortality. All-cause death was observed in 168 patients and MACEs in 343 patients during follow up period (median 387 days, interquartile range; 239 - 727 days). There was a significant difference in all-cause death (Left; p=0.0003 by log-rank test) and MACEs (Rights; p=0.007 by log-rank test) among the 4 groups based on the quartile of the TyG index, and the highest TyG group (Q4) had lower incidence of all-cause death and MACEs. * p<0.005, † p<0.001 vs. Q1; ‡ p<0.05 vs. Q2.

**Table 3.**
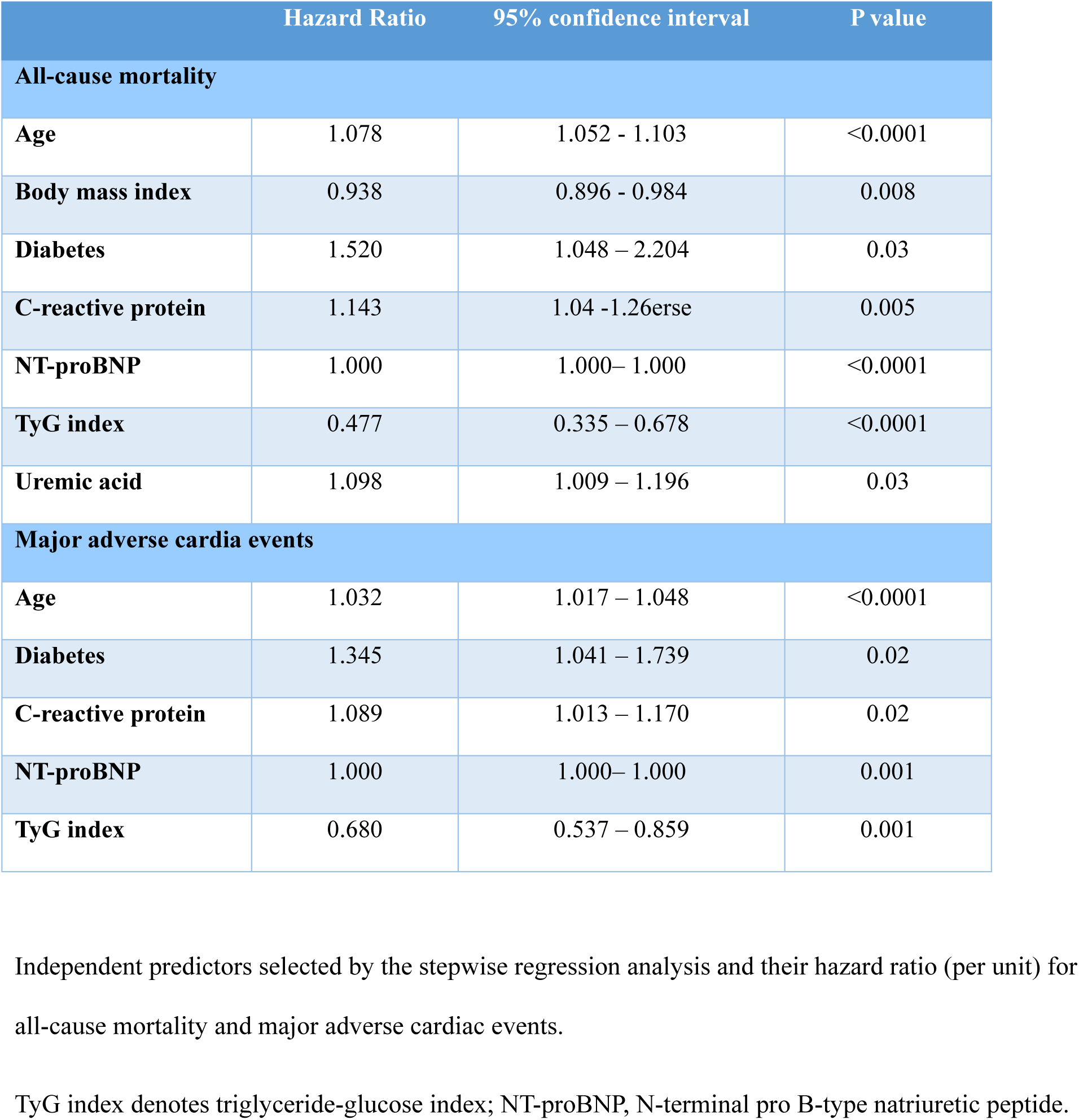
Results of stepwise regression analysis.

|  | Hazard Ratio | 95% confidence interval | P value |
| --- | --- | --- | --- |
| **All-cause mortality** |  |  |  |
| **Age** | 1.078 | 1.052 - 1.103 | <0.0001 |
| **Body mass index** | 0.938 | 0.896 - 0.984 | 0.008 |
| **Diabetes** | 1.520 | 1.048 – 2.204 | 0.03 |
| **C-reactive protein** | 1.143 | 1.04 - 1.26 | 0.005 |
| **NT-proBNP** | 1.000 | 1.000– 1.000 | <0.0001 |
| **TyG index** | 0.477 | 0.335 – 0.678 | <0.0001 |
| **Uremic acid** | 1.098 | 1.009 – 1.196 | 0.03 |
| **Major adverse cardia events** |  |  |  |
| **Age** | 1.032 | 1.017 – 1.048 | <0.0001 |
| **Diabetes** | 1.345 | 1.041 – 1.739 | 0.02 |
| **C-reactive protein** | 1.089 | 1.013 – 1.170 | 0.02 |
| **NT-proBNP** | 1.000 | 1.000– 1.000 | 0.001 |
| **TyG index** | 0.680 | 0.537 – 0.859 | 0.001 |

Independent predictors selected by the stepwise regression analysis and their hazard ratio (per unit) for all-cause mortality and major adverse cardiac events.
TyG index denotes triglyceride-glucose index; NT-proBNP, N-terminal pro B-type natriuretic peptide.

There was no significant difference in HR for all-cause death or for MACEs between subgroups based on age, sex, BMI, diabetes, HbA1c, history of hypertension, dyslipidemia, chronic kidney disease and atrial fibrillation (Figure 4).

**Figure 4.**
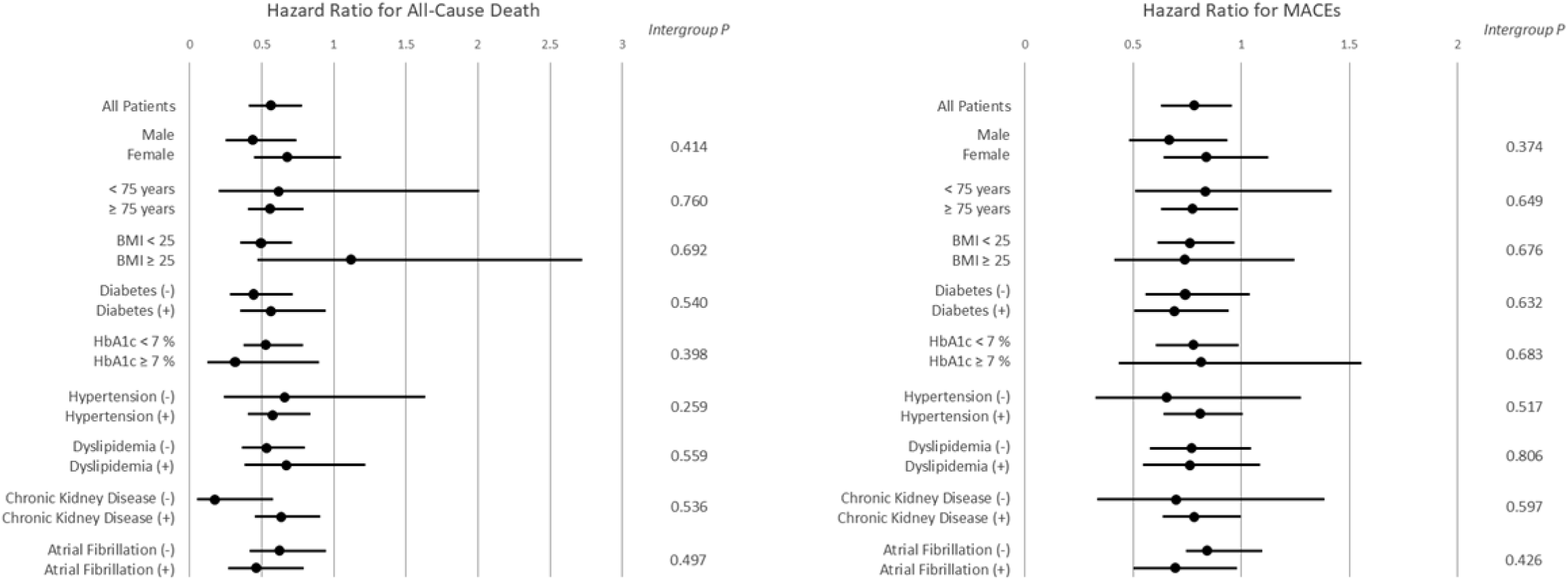
Subgroup analysis of hazard ratio per unit of the TyG index. There was no difference in hazard ratio (HR) for all-cause death (Left) or for MACEs (Right) between subgroups based on age, sex, BMI, diabetes, HbA1c, history of hypertension, dyslipidemia, chronic kidney disease and atrial fibrillation.

## Discussion

We investigated the association of the TyG index and clinical outcomes in 917 patients hospitalized for acute decompensated HFpEF, in which 39.2% of them had diabetes. Counterintuitively, a stepwise Cox proportional hazards model revealed that a higher TyG index was independently associated with a lower risk of both all-cause death (HR 0.53) and MACEs (HR 0.77) during follow-up period (median 387 days). When patients were stratified into quartiles based on the TyG index, Kaplan-Meier survival curves demonstrated a lower cumulative incidence of both endpoints in the higher TyG index groups. This paradoxical association between the TyG index and prognosis remained consistent across various subgroups, including diabetes status, age, sex, BMI, and HbA1c and other risk factors. No significant differences were observed in echocardiographic parameters except that the lowest TyG group had a lower E/e’ ratio but conversely a greater LAVI compared to the higher TyG groups, implying that IR had little effect on systolic/diastolic cardiac function. These results suggested that, whereas IR generally contributes to the development of HF, IR may be associated with better clinical outcomes in this cohort of patients with established HFpEF.

### TyG index as an index of IR

While IR is frequently observed in patients with incident heart HF irrespective of presence or absence of diabetes^3,4,16^, the association between IR and prognosis in patients with established HF is controversial and inconclusive. HOMA-IR was associated with death or readmission at 90 days but not at 1 year in patients hospitalized for HF in which half of them had EF>45%^17^. Other studies investigated the correlation of the TyG index with clinical outcomes in HF patients^18,19^. Zhou et al. reported that patients in the highest tertile of the TyG index (≥9.32)) had higher rate of all cause- and cardiovascular death than those in the lowest tertile (<8.83) in 823 patients with HFpEF, whereas no difference was observed in readmission rate^18^. The difference between their results and present ones may be explained by the different clinical characteristics of the study cohorts.

The median age of 83 years in the present study was far older than most of previous large scale HF studies; the mean age of HFpEF patients in the Practice Innovation And Clinical Excellence (PINNACLE) registry was 69.7 ± 13.6 years^20^, and that of recent prospective studies for HFpEF was around 70 years^21–24^. On the other hand, patients in the large scale, Japanese HF registries were as old as those in the present cohort. The mean age of the nationwide Japanese Registry Of Acute Decompensated Heart Failure (JROADHF) was 78.0±12.5 years and 68.9% of them was older than 75 years^25^. Another nationwide, Japanese Heart Failure Syndrome With Preserved Ejection Fraction (JASPER) Registry had median age of 80 years^26^. The mean age in the study of Zhou et al. was 73.0±12.7 years, which may be related with the different results between two studies^18^.

BMI was another major difference between the present and other studies. The median BMI was only 21.5 kg/m^2^, and even that of highest quartile of the TyG index (Q4) was 23.2 kg/m^2^. Patients with BMI ≥25.0 kg/m^2^ were only 20.9% while those with BMI<18.5 kg/m^2^ accounted for 20.2% of the patients. BMI in the large scale registry and prospective HFpEF studies was around 30 kg/m^2 20–24,27^. BMI tended to decrease with increasing age, but even patients ≥ 75 years in the DELIVER trial had BMI of 28.7±5.5 kg/m^2 24^. BMI in Japanese HFpEF registries was similar to the present study (JROADHF, 22.9±4.6 kg/m^2^ and JASPER, 23.9±4.7 kg/m^2^)^25,26^.

The mean BMI in the study of Zhou et al. was 25.5±4.7 kg/m^2^ and even than of the lowest tertile of the TyG index was 24.6±4.6 kg/m^2 18^, which was higher than that in the present study. The small BMI in the present study led to lower TyG index (8.54±0.53, range 7.26 to 10.60) compared to other studies. The J-shape relation between the TyG index and all cause- or cardiovascular death in HFpEF was observed in the previous study, and nadir of mortality was present around 8.5 of the TyG index^18^. Half of the present study patients (Q1+Q2) had the TyG index below it, which may be one of the reasons for the different results between two studies.

The cohort in the present study was predominantly old and had a low BMI, which may be consistent with the trend in patients with HFpEF in Japan, the world’s most rapidly aging society. An analysis on the characteristics of Japanese patients with hospitalized for HFpEF revealed that old and low BMI patients with some comorbidities were predominant and that those with high BMI and left ventricular hypertrophy was only 10% of patients ^28^. This specific profile of HFpEF patients in Japan may be related to the paradoxical association between the TyG index and prognosis. Although the beneficial effect of IR was observed across age and BMI in the present study patients, it is unclear whether similar effect could be present among younger and more obese HFpEF patients in other populations. Recent studies demonstrated that glucagon-like peptide-1 receptor (GLP-1) agonists or glucose-dependent insulinotropic polypeptide (GIP)/GLP-1 receptor agonists improve clinical manifestation and outcomes in obese patients with HFpEF regardless of diabetes status^29,30^, and body weight reduction and anti-inflammatory action is considered as major mechanisms of these effects^31,32^. The beneficial effects of IR may be blunted by the detrimental effects of overweight in obese patients.

### Clinical Implications

The obesity paradox in HF is a complex and intriguing phenomenon and its presence is still controversial^33–35^. Whereas weight loss improves quality of life and prognosis in HF patients with overweight and obesity^36^, it is associated with poor clinical prognosis in those without high obesity, especially in lean patients^37,38^. The underlying mechanisms of the obesity paradox are not fully understood, and the interplay between body and fat composition, cardiorespiratory fitness, and metabolic factors may contribute to this phenomenon. The paradoxical effects of IR observed in this study may contribute to the obesity paradox in HFpEF patients without obesity.

Thiazolidinediones are effective at improving IR in diabetic patients but they are associated with an increased risk of HF and prescribing them must be cautious for HF patints^39–41^. The mechanisms of the increased HF risk are complex and may involve several factors. Thiazolidinediones can cause fluid retention mostly through sodium retention in the distal nephron^42,43^, leading to increase in cardiac preload and potentially afterload. Other possible mechanisms of fluid retention include sympathetic activation, increased vascular permeability, and the response to vasodilatation^41^. The reduction of beneficial effect of IR observed in this study also may contribute to the deleterious effect of thiazolidinediones.

### Limitation of the study

The present study was a retrospective study from a registered database, and the number of study patients was limited. The TyG index could not be calculated in 178 patients (16.3 %), of which 17 patients died before discharge and 167 patients had missing parameters for the calculation of the index, which may introduce some selection bias. The TyG index was determined only at discharge, and it may change during the follow-up period by treatment and life-style modification. Regional or ethnic differences exist in the mortality and morbidity of HFpEF^44,45^, which may be related with the different patient characteristics as described above. Moreover, the effectiveness of the TyG index to predict cardiovascular disease and mortality varies across different ethnic groups^46^. Thus, it is unclear whether the present results are applicable to patients outside East Asia.

Despite these study limitations, the present study provided new insights into the complex relationship between IR and clinical outcomes in patients with established HFpEF.

## Data Availability

The datasets generated and/or analyzed during the current study are available from the corresponding author on reasonable request.

## Acknowledgements

The authors thank all PURSUIT-HFpEF study participants whose names are listed in the Supplementary Appendix for their support of the present study.

## References

1. Ndumele CE, Rangaswami J, Chow SL, Neeland IJ, Tuttle KR, Khan SS, Coresh J, Mathew RO, Baker-Smith CM, Carnethon MR, et al, on behalf of the American Heart Association. Cardiovascular-Kidney-Metabolic Health: A Presidential Advisory From the American Heart Association. Circulation. 2023;148:1606–1635.

2. Wamil M, Coleman RL, Adler AI, McMurray JJV, Holman RR. Increased Risk of Incident Heart Failure and Death Is Associated With Insulin Resistance in People With Newly Diagnosed Type 2 Diabetes: UKPDS 89. Diabetes Care. 2021;44:1877–1884.

3. Vardeny O, Gupta DK, Claggett B, Burke S, Shah A, Loehr L, Rasmussen-Torvik L, Selvin E, Chang PP, Aguilar D, et al. Insulin Resistance and Incident Heart Failure. JACC Heart Fail. 2013;1:531–536.

4. Banerjee D, Biggs ML, Mercer L, Mukamal K, Kaplan R, Barzilay J, Kuller L, Kizer JR, Djousse L, Tracy R, et al. Insulin Resistance and Risk of Incident Heart Failure: Cardiovascular Health Study. Circ Heart Fail. 2013;6:364–370.

5. Erqou S, Adler AI, Challa AA, Fonarow GC, Echouffo-Tcheugui JB. Insulin resistance and incident heart failure: a META-ANALYSIS. Eur J Heart Fail. 2022;24:1139–1141.

6. Gudenkauf B, Shaya G, Mukherjee M, Michos ED, Madrazo J, Mathews L, Shah SJ, Sharma K, Hays AG. Insulin resistance is associated with subclinical myocardial dysfunction and reduced functional capacity in heart failure with preserved ejection fraction. J Cardiol. 2024;83:100–104.

7. Son TK, Toan NH, Thang N, Le Trong Tuong H, Tien HA, Thuy NH, Van Minh H, Valensi P. Prediabetes and insulin resistance in a population of patients with heart failure and reduced or preserved ejection fraction but without diabetes, overweight or hypertension. Cardiovasc Diabetol. 2022;21:75.

8. Scherbakov N, Bauer M, Sandek A, Szabó T, Töpper A, Jankowska EA, Springer J, Von Haehling S, Anker SD, Lainscak M, et al. Insulin resistance in heart failure: differences between patients with reduced and preserved left ventricular ejection fraction. Eur J Heart Fail. 2015;17:1015–1021.

9. Guerrero-Romero F, Simental-Mendía LE, González-Ortiz M, Martínez-Abundis E, Ramos-Zavala MG, Hernández-González SO, Jacques-Camarena O, Rodríguez-Morán M. The product of triglycerides and glucose, a simple measure of insulin sensitivity. Comparison with the euglycemic-hyperinsulinemic clamp. J Clin Endocrinol Metab. 2010;95:3347–3351.

10. Wan H, Cao H, Ning P. Superiority of the triglyceride glucose index over the homeostasis model in predicting metabolic syndrome based on NHANES data analysis. Sci Rep. 2024;14:15499.

11. Son D-H, Lee HS, Lee Y-J, Lee J-H, Han J-H. Comparison of triglyceride-glucose index and HOMA-IR for predicting prevalence and incidence of metabolic syndrome. Nutr Metab Cardiovasc Dis. 2022;32:596–604.

12. Fang Y, Shen J, Lyu L. Value of the triglyceride–glucose index and related parameters in heart failure patients. Front Cardiovasc Med. 2024;11:1397907.

13. Khalaji A, Behnoush AH, Khanmohammadi S, Ghanbari Mardasi K, Sharifkashani S, Sahebkar A, Vinciguerra C, Cannavo A. Triglyceride-glucose index and heart failure: a systematic review and meta-analysis. Cardiovasc Diabetol. 2023;22:244.

14. Xu L, Wu M, Chen S, Yang Y, Wang Y, Wu S, Tian Y. Triglyceride–glucose index associates with incident heart failure: A cohort study. Diabetes Metab. 2022;48:101365.

15. Suna S, Hikoso S, Yamada T, Uematsu M, Yasumura Y, Nakagawa A, Takeda T, Kojima T, Kida H, Oeun B, et al. Study protocol for the PURSUIT-HFpEF study: a Prospective, Multicenter, Observational Study of Patients with Heart Failure with Preserved Ejection Fraction. BMJ Open. 2020;10:e038294.

16. Ingelsson E, Sundström J, Ärnlöv J, Zethelius B, Lind L. Insulin Resistance and Risk of Congestive Heart Failure. JAMA. 2005;294:334.

17. Castillo Costa Y, Mauro V, Fairman E, Charask A, Olguín L, Cáceres L, Barrero C. Prognostic Value of Insulin Resistance Assessed by HOMA-IR in Non-Diabetic Patients with Decompensated Heart Failure. Curr Probl Cardiol. 2023;48:101112.

18. Zhou Q, Yang J, Tang H, Guo Z, Dong W, Wang Y, Meng X, Zhang K, Wang W, Shao C, et al. High triglyceride-glucose (TyG) index is associated with poor prognosis of heart failure with preserved ejection fraction. Cardiovasc Diabetol. 2023;22:263.

19. Huang R, Wang Z, Chen J, Bao X, Xu N, Guo S, Gu R, Wang W, Wei Z, Wang L. Prognostic value of triglyceride glucose (TyG) index in patients with acute decompensated heart failure. Cardiovasc Diabetol. 2022;21:88.

20. Ibrahim NE, Song Y, Cannon CP, Doros G, Russo P, Ponirakis A, Alexanian C, Januzzi Jr JL. Heart failure with mid-range ejection fraction: characterization of patients from the PINNACLE Registry®. ESC Heart Fail. 2019;6:784–792.

21. Shah SJ, Heitner JF, Sweitzer NK, Anand IS, Kim H-Y, Harty B, Boineau R, Clausell N, Desai AS, Diaz R, et al. Baseline Characteristics of Patients in the Treatment of Preserved Cardiac Function Heart Failure With an Aldosterone Antagonist Trial. Circ Heart Fail. 2013;6:184–192.

22. Solomon SD, Rizkala AR, Lefkowitz MP, Shi VC, Gong J, Anavekar N, Anker SD, Arango JL, Arenas JL, Atar D, et al. Baseline Characteristics of Patients With Heart Failure and Preserved Ejection Fraction in the PARAGON-HF Trial. Circ Heart Fail. 2018;11:e004962.

23. Anker SD, Butler J, Filippatos G, Shahzeb Khan M, Ferreira JP, Bocchi E, Böhm M, Brunner-La Rocca HP, Choi D, Chopra V, et al. EMPEROR-Preserved Trial Committees and Investigators. Baseline characteristics of patients with heart failure with preserved ejection fraction in the EMPEROR-Preserved trial. Eur J Heart Fail. 2020;22:2383–2392.

24. Peikert A, Martinez FA, Vaduganathan M, Claggett BL, Kulac IJ, Desai AS, Jhund PS, De Boer RA, DeMets D, Hernandez AF, et al. Efficacy and Safety of Dapagliflozin in Heart Failure With Mildly Reduced or Preserved Ejection Fraction According to Age: The DELIVER Trial. Circ Heart Fail 2022;15:e010080.

25. Ide T, Kaku H, Matsushima S, Tohyama T, Enzan N, Funakoshi K, Sumita Y, Nakai M, Nishimura K, Miyamoto Y, et al, the JROADHF Investigators. Clinical Characteristics and Outcomes of Hospitalized Patients With Heart Failure From the Large-Scale Japanese Registry Of Acute Decompensated Heart Failure (JROADHF). Circ J. 2021;85:1438–1450.

26. Nagai T, Yoshikawa T, Saito Y, Takeishi Y, Yamamoto K, Ogawa H, Anzai T, on behalf of the JASPER Investigators. Clinical Characteristics, Management, and Outcomes of Japanese Patients Hospitalized for Heart Failure With Preserved Ejection Fraction ― A Report From the Japanese Heart Failure Syndrome With Preserved Ejection Fraction (JASPER) Registry ―. Circ J. 2018;82:1534–1545.

27. Adamson C, Kondo T, Jhund PS, De Boer RA, Cabrera Honorio JW, Claggett B, Desai AS, Alcocer Gamba MA, Al Habeeb W, Hernandez AF, Inzucchi SE,et al. Dapagliflozin for heart failure according to body mass index: the DELIVER trial. Eur Heart J. 2022;43:4406–4417.

28. Kyodo A, Kanaoka K, Keshi A, Nogi M, Nogi K, Ishihara S, Kamon D, Hashimoto Y, Nakada Y, Ueda T, et al. Heart failure with preserved ejection fraction phenogroup classification using machine learning. ESC Heart Fail. 2023;10:2019–2030.

29. Kosiborod MN, Abildstrøm SZ, Borlaug BA, Butler J, Rasmussen S, Davies M, Hovingh GK, Kitzman DW, Lindegaard ML, Møller DV, et al. Semaglutide in Patients with Heart Failure with Preserved Ejection Fraction and Obesity. N Engl J Med. 2023;389:1069–1084.

30. Packer M, Zile MR, Kramer CM, Baum SJ, Litwin SE, Menon V, Ge J, Weerakkody GJ, Ou Y, Bunck MC, et al. Tirzepatide for Heart Failure with Preserved Ejection Fraction and Obesity. N Engl J Med. 2025;392:427–437.

31. Borlaug BA, Zile MR, Kramer CM, Baum SJ, Hurt K, Litwin SE, Murakami M, Ou Y, Upadhyay N, Packer M. Effects of tirzepatide on circulatory overload and end-organ damage in heart failure with preserved ejection fraction and obesity: a secondary analysis of the SUMMIT trial. Nat Med. 2025;31:544–551.

32. Borlaug BA, Kitzman DW, Davies MJ, Rasmussen S, Barros E, Butler J, Einfeldt MN, Hovingh GK, Møller DV, Petrie MC, et al. Semaglutide in HFpEF across obesity class and by body weight reduction: a prespecified analysis of the STEP-HFpEF trial. Nat Med. 2023;29:2358–2365.

33. Butt JH, Petrie MC, Jhund PS, Sattar N, Desai AS, Køber L, Rouleau JL, Swedberg K, Zile MR, Solomon SD, et al. Anthropometric measures and adverse outcomes in heart failure with reduced ejection fraction: revisiting the obesity paradox. Eur Heart J. 2023;44:1136–1153.

34. Butt JH, McMurray JJV. Obesity paradox can be a fact: unveiling the hidden role of adipose tissue. Eur Heart J. 2024;45:2168–2168.

35. Zhuang C, Chen Y, Ruan J, Yu H, Zhu P, Zhu Y. Correlation between the prognostic nutritional index and outcomes in older patients aged ≥ 60 years with chronic heart failure. Int J Clin Pharm. 2023;45:163–173.

36. Chi M, Nie Y, Su Y, Wang N, Li A, Ma T, Hou Y. Effects of weight loss in heart failure patients with overweight and obesity: a systematic review and meta-analysis. Eur J Prev Cardiol. 2023;30:1906–1921.

37. Wu X, Wang Y, Hu X. Association of weight loss with cardiovascular or all-cause mortality in patients with heart failure: A meta-analysis. Int J Obes. 2024;48:626–634.

38. Pocock SJ, McMurray JJV, Dobson J, Yusuf S, Granger CB, Michelson EL, Ostergren J, Pfeffer MA, Solomon SD, Anker SD, et al, on behalf of the CHARM Investigators. Weight loss and mortality risk in patients with chronic heart failure in the candesartan in heart failure: assessment of reduction in mortality and morbidity (CHARM) programme. Eur Heart J. 2008;29:2641–2650.

39. Erdmann E, Wilcox RG. Weighing up the cardiovascular benefits of thiazolidinedione therapy: the impact of increased risk of heart failure. Eur Heart J. 2007;29:12–20.

40. Kaul S, Bolger AF, Herrington D, Giugliano RP, Eckel RH. Thiazolidinedione Drugs and Cardiovascular Risks: A Science Advisory From the American Heart Association and American College of Cardiology Foundation. Circulation. 2010;121:1868–1877.

41. Nesto RW, Bell D, Bonow RO, Fonseca V, Grundy SM, Horton ES, Le Winter M, Porte D, Semenkovich CF, Smith S, et al, American Heart Association, American Diabetes Association. Thiazolidinedione use, fluid retention, and congestive heart failure: a consensus statement from the American Heart Association and American Diabetes Association. October 7, 2003. Circulation. 2003;108:2941–2948.

42. Zhang H, Zhang A, Kohan DE, Nelson RD, Gonzalez FJ, Yang T. Collecting duct-specific deletion of peroxisome proliferator-activated receptor gamma blocks thiazolidinedione-induced fluid retention. Proc Natl Acad Sci U S A. 2005;102:9406–9411.

43. Guan Y, Hao C, Cha DR, Rao R, Lu W, Kohan DE, Magnuson MA, Redha R, Zhang Y, Breyer MD. Thiazolidinediones expand body fluid volume through PPARgamma stimulation of ENaC-mediated renal salt absorption. Nat Med. 2005;11:861–866.

44. Pandey A, Omar W, Ayers C, LaMonte M, Klein L, Allen NB, Kuller LH, Greenland P, Eaton CB, Gottdiener JS, et al. Sex and Race Differences in Lifetime Risk of Heart Failure With Preserved Ejection Fraction and Heart Failure With Reduced Ejection Fraction. Circulation. 2018;137:1814–1823.

45. Ziaeian B, Heidenreich PA, Xu H, DeVore AD, Matsouaka RA, Hernandez AF, Bhatt DL, Yancy CW, Fonarow GC. Race/Ethnic Differences in Outcomes Among Hospitalized Medicare Patients With Heart Failure and Preserved Ejection Fraction. JACC Heart Fail. 2017;5:483–493.

46. Lopez-Jaramillo P, Gomez-Arbelaez D, Martinez-Bello D, Abat MEM, Alhabib KF, Avezum Á, Barbarash O, Chifamba J, Diaz ML, Gulec S, et al. Association of the triglyceride glucose index as a measure of insulin resistance with mortality and cardiovascular disease in populations from five continents (PURE study): a prospective cohort study. Lancet Healthy Longev. 2023;4:e23–e33.

